# A vendor-neutral functional MRI acquisition protocol for multi-site studies

**DOI:** 10.1101/2025.08.27.25334579

**Authors:** Jon-Fredrik Nielsen, Maximillian K. Egan, Qingping Chen, Mojtaba Shafiekhani, Jeffrey S. Soldate, Jonathan Lisinksi, Stephen M. LaConte, Bradley P. Sutton, R. Allen Waggoner, Jeffrey A. Fessler, Douglas C. Noll, Maxim Zaitsev, Scott J. Peltier

## Abstract

We present an open, vendor-neutral BOLD SMS-EPI protocol tailored for multi-site fMRI studies, intended as a drop-in replacement for conventional vendor-specific, black-box acquisition and reconstruction pipelines. Built on Pulseq—an emerging standard for cross-platform MRI pulse sequence development—our protocol ensures identical SMS-EPI pulse sequences and image reconstruction across scanner vendors. This provides, for the first time, known and consistent experimental conditions across sites and scanner software versions.

We begin by reviewing the current capabilities of the Pulseq framework, including vendor support and safety considerations. We then detail our SMS-EPI implementation and demonstrate its performance using resting-state fMRI pilot data from healthy volunteers, showing reduced site variance compared to corresponding vendor protocols on Siemens and GE scanners.

To support adoption, we provide practical resources to help researchers integrate Pulseq fMRI into their studies, including example text for grant proposals and IRB submissions. These resources are freely available at https://github.com/HarmonizedMRI/Functional.

Our vision is for Pulseq fMRI to become the standard for multi-site research, enabling more reproducible science and serving as a reference for the development of novel acquisition and reconstruction methods.

## Introduction

Pooling functional MRI (fMRI) data across multiple sites can enhance statistical power and support the inclusion of more diverse and representative subject populations. However, these benefits can only be fully realized if acquisition protocols are standardized, transparent, and reproducible, thereby not introducing additional variance to the data. In current practice, most fMRI workflows depend on vendor-specific, opaque (“black-box”) acquisition and image reconstruction pipelines. However, the scanner software is typically optimized for operator convenience rather than scientific rigor, making the protocols based on vendor-provided sequences non-portable, difficult to replicate, and poorly suited for cross-site harmonization.

As a result, the stability of the fMRI image formation process cannot be guaranteed across scanners from different MRI vendors—or even across different software versions on the same scanner. For example, Keenan et al.^1^ observed inconsistent flip angles (RF transmit amplitude) following a scanner software upgrade. Consequently, observed differences in functional measures between sites or over time may reflect systematic differences in protocol implementation rather than genuine biological variation. This lack of reproducibility also creates a barrier to innovation, as implementing new fMRI acquisition strategies within proprietary vendor environments is technically demanding and time-consuming.

This article describes a vendor-neutral, open-source, and fully harmonized fMRI protocol that addresses these challenges. Our acquisition protocol is implemented in Pulseq^2^, an emerging open standard for describing MRI pulse sequences in a vendor-independent way. This allows for *exact* harmonization of the pulse sequence across vendor platforms down to the individual RF/gradient waveform level, which in turn ensures that important aspects of the acquisition, such as the nominal slice profile and choice of fat suppression method, are kept constant across sites and over time. Equally important, Pulseq enables harmonization of the image reconstruction step: since the raw (k-space) data and coil calibration data are acquired in a known way, the image time-series can be reconstructed in exactly the same way regardless of which scanner it came from. This stands in contrast to vendor-specific product protocols, which often employ different—and typically opaque—techniques for parallel imaging (e.g., SENSE^3^, GRAPPA^4^, or deep learning-based methods) and image pre-processing. Pulseq therefore creates an opportunity for the fMRI community to standardize not only the pulse sequence, but also the image reconstruction algorithms and associated scanner and coil calibration scans.

We begin by outlining the **fundamentals and current capabilities of the Pulseq** framework, including how it enables rapid prototyping and deployment of custom MRI sequences across different vendor platforms, and ensures immunity to scanner software upgrades. This section is primarily intended for MRI pulse sequence programmers and protocol maintainers.

We then describe the implementation of **our Pulseq fMRI protocol** and the practical steps needed to run the sequence on two major MRI vendor platforms—Siemens and GE. We also present **traveling subject data** that illustrate the feasibility of deploying our protocol in a multi-site research setting. Notably, these results show reduced cross-vendor variability in functional connectivity measures compared to standard vendor-specific (product) protocols.

Finally, we provide **recommendations and additional resources for fMRI researchers** seeking to incorporate Pulseq into their own experimental protocols and grant applications.

### The Pulseq platform

The key technical component of our open-source protocol relates to the initial part of the image formation chain: the signal generation and the data acquisition steps, where we employ the Pulseq platform for vendor-neutral MRI pulse sequence programming. Pulseq is both a conceptual sequence representation, a file specification that encapsulates it, and a set of software tools for creating, analyzing, and executing sequence files. Here we attempt to clarify these concepts and provide the MRI physicist or protocol maintainer with enough information to install and run Pulseq sequences on their scanner.

#### Overall workflow

The key idea behind Pulseq is to write the MRI pulse sequence specification to a vendor-neutral file that is to be loaded onto a particular MRI scanner via a vendor-specific interpreter (Fig. 1). That same Pulseq file can also be used for other purposes, such as performing Bloch simulations, and generating sequence timing diagrams or precise k-space trajectories. The Pulseq standard itself is vendor-neutral and agnostic to the specifics of a particular vendor platform and the implementation details of the specific interpreter.

**Figure 1.**
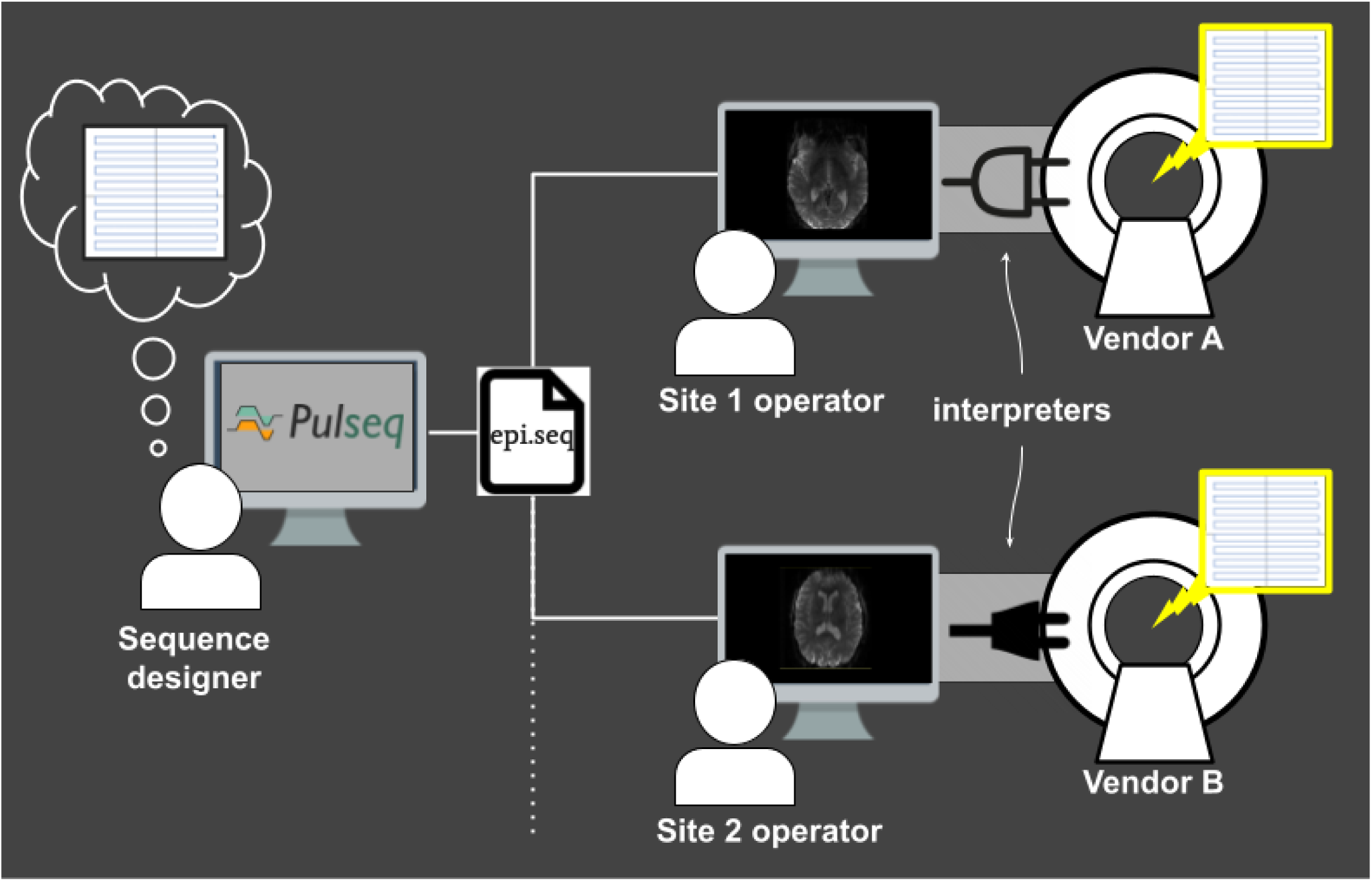
Overview of the Pulseq platform for implementing and sharing vendor-neutral pulse sequences. The sequence designer creates a seq file that specifies all low-level details of the sequence that she envisions. That .seq file is shared with scanner operators at different sites. At each site, a vendor-specific and sequence-agnostic interpreter executable loads and executes the sequence on the scanner. This workflow ensures perfect harmonization of the pulse sequence across sites and is easy to maintain across scanner system upgrades since only the interpreter needs to be recompiled. Computer screen image downloaded from https://commons.wikimedia.org/.

Once the Pulseq interpreter is installed on a particular scanner, different pulse sequences can be executed by simply selecting the appropriate sequence file from the user interface of the scanner console; the interpreter itself is sequence-agnostic. Importantly, maintaining Pulseq sequences across system software upgrades is simply a matter of recompiling the interpreter, since the sequence files are agnostic to the scanner software version. This greatly simplifies the task of maintaining fMRI protocols across scanner software versions.

### A deeper dive into Pulseq

While this article focuses on a specific SMS-EPI fMRI protocol developed and validated by the authors, researchers across the broader fMRI community may want to modify the sequence to suit their own needs. Whether modifying an existing Pulseq sequence or creating one from scratch, it is important to understand how Pulseq represents the various elements of the sequence, what software tools are available to implement them, and how to ensure compatibility with a particular scanner make and model. This section is intended to support that understanding. However, readers who are primarily interested in using the pre-configured fMRI sequence files provided by the authors may choose to skip it.

#### Sequence representation

In Pulseq, an MRI pulse sequence is represented as a series of sequential, non-overlapping blocks (Fig. 2). Each block can contain at most one radiofrequency (RF) pulse, one gradient waveform per axis, and one data acquisition (ADC) window. Apart from this constraint, and the usual hardware limits on peak waveform amplitude and gradient slew rate, the sequence designer is free to choose where to place the block boundaries. The sequence is defined explicitly, without formal relationships between blocks or higher-level constructs such as loops or defined repetition time (TR) intervals.

**Figure 2.**
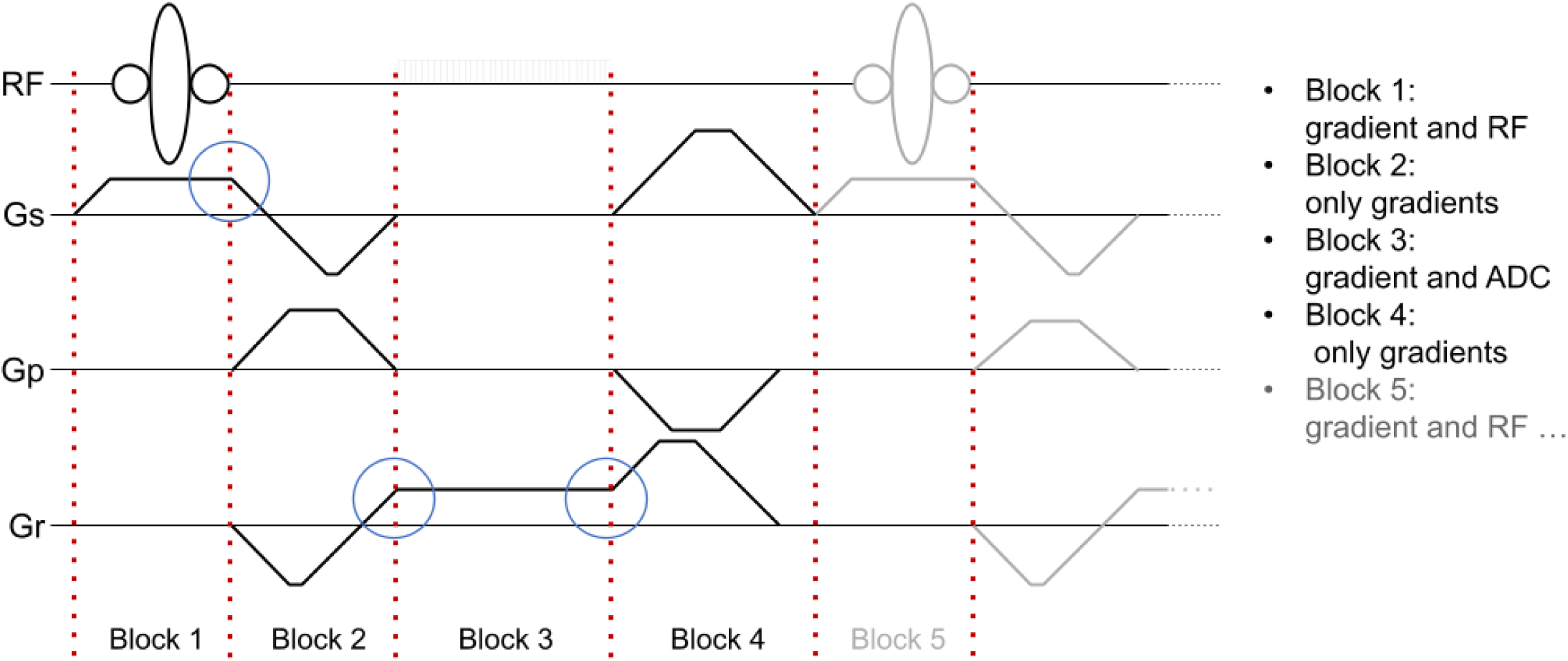
Pulseq sequence representation. Non-overlapping blocks are executed consecutively and without gaps. Each waveform channel can contain at most one event within a block. Gradient waveforms must be continuous across block boundaries (blue circles). RF: radiofrequency; Gs: gradient along slice-select (“z”) axis; Gp: gradient along phase-encode (y) axis; Gr: gradient along readout (x) axis.

#### Defining sequence events

The core task for a Pulseq sequence programmer is to define the RF, gradient, and ADC events that make up the sequence. Each event is described using a small set of parameters grouped into a common namespace (e.g., an object or struct). For instance, an RF event must specify its start time relative to the beginning of the block (delay), the complex waveform samples in physical units (signal) and their corresponding time points (t), the frequency offset in Hz (freqOffset) or in parts-per-million (ppmOffset), and the phase offset in radians (phaseOffset). Additionally, system-specific parameters such as deadTime and ringdownTime, which reflect RF coil and amplifier switching behavior, affect the timing of the RF events. The sequence designer can either manually construct this RF event struct or use the helper functions provided in the Pulseq MATLAB (Fig. 3) or Python libraries^5,6^. The same approach applies to gradient and ADC events that can be created using library functions or by manually specifying the required parameters.

**Figure 3.**
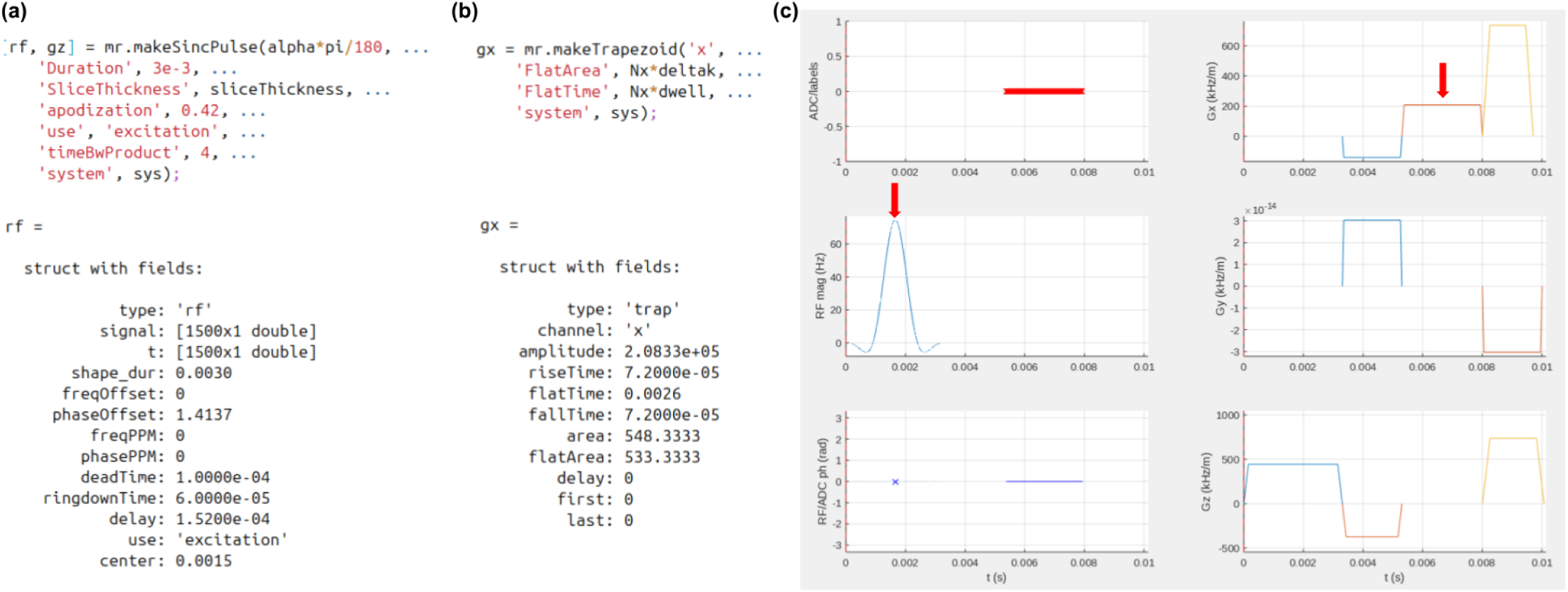
Examples of (a) RF and (b) gradient event creation using the Pulseq MATLAB toolbox. Once the RF or gradient struct is created, it can be freely modified if desired; it is a simple struct data type, (c) Sequence timing diagram showing the location of these RF and gradient events (red arrows) in a gradient echo sequence.

**Figure 4.**
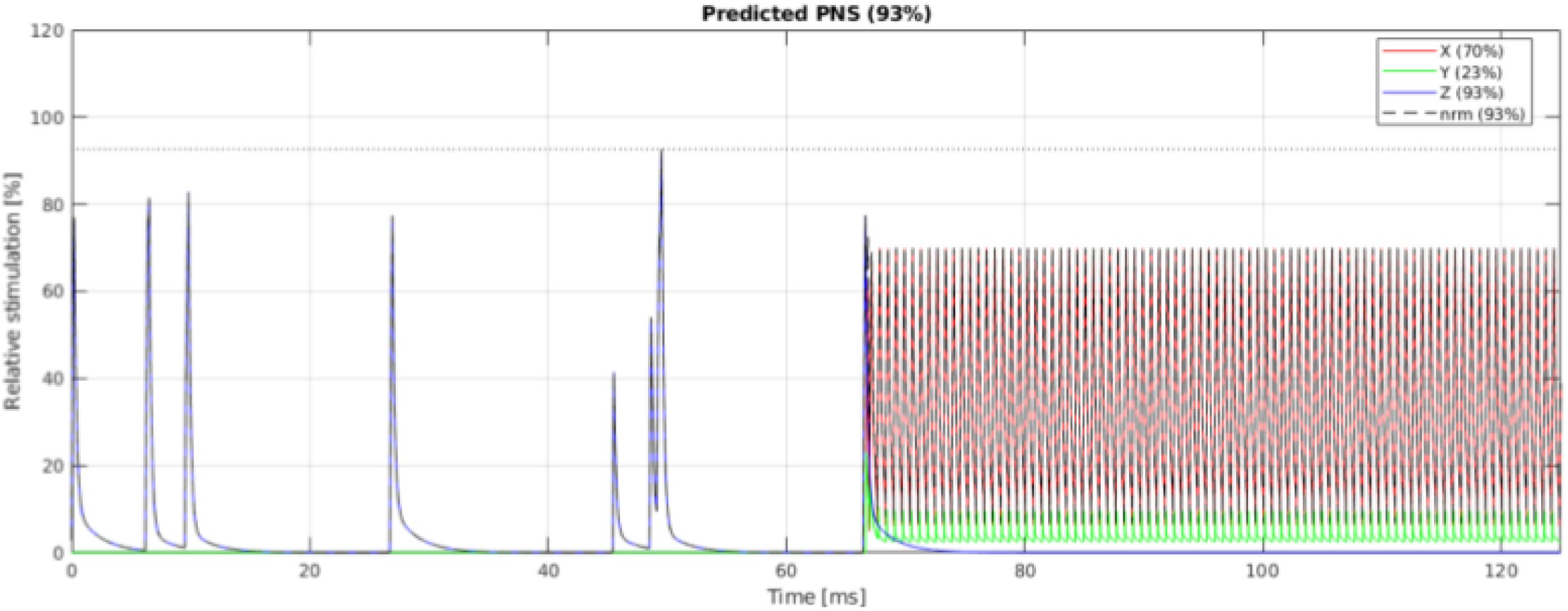
Example of PNS prediction in MATLAB for a Siemens PRISMA scanner using the SAFE^16^ model implementation by Szczepankiewicz et al.^15,17^. Such modeling can be done prior to executing the sequence on the scanner, as a step toward ensuring patient safety.

#### Assembling the sequence

Once the individual events have been defined, they need to be assigned to blocks and added to a sequence object. This process typically begins by creating an empty sequence object that is configured for a particular scanner hardware platform, followed by inserting blocks into that sequence object using the addBlock() member function. Blocks are added in the order of execution, and the Pulseq toolbox automatically manages the internal organization of the sequence for you. Although the Pulseq representation does not formally define relationships between blocks, it is common practice to use a loop structure when assembling the sequence in code. Examples of MATLAB and Python code for creating many basic and advanced sequences are freely available on Github; some of these sites are listed in Table 1. Once the sequence object is populated with blocks, it can be plotted for visual inspection and written to a file.

**Table 1.**
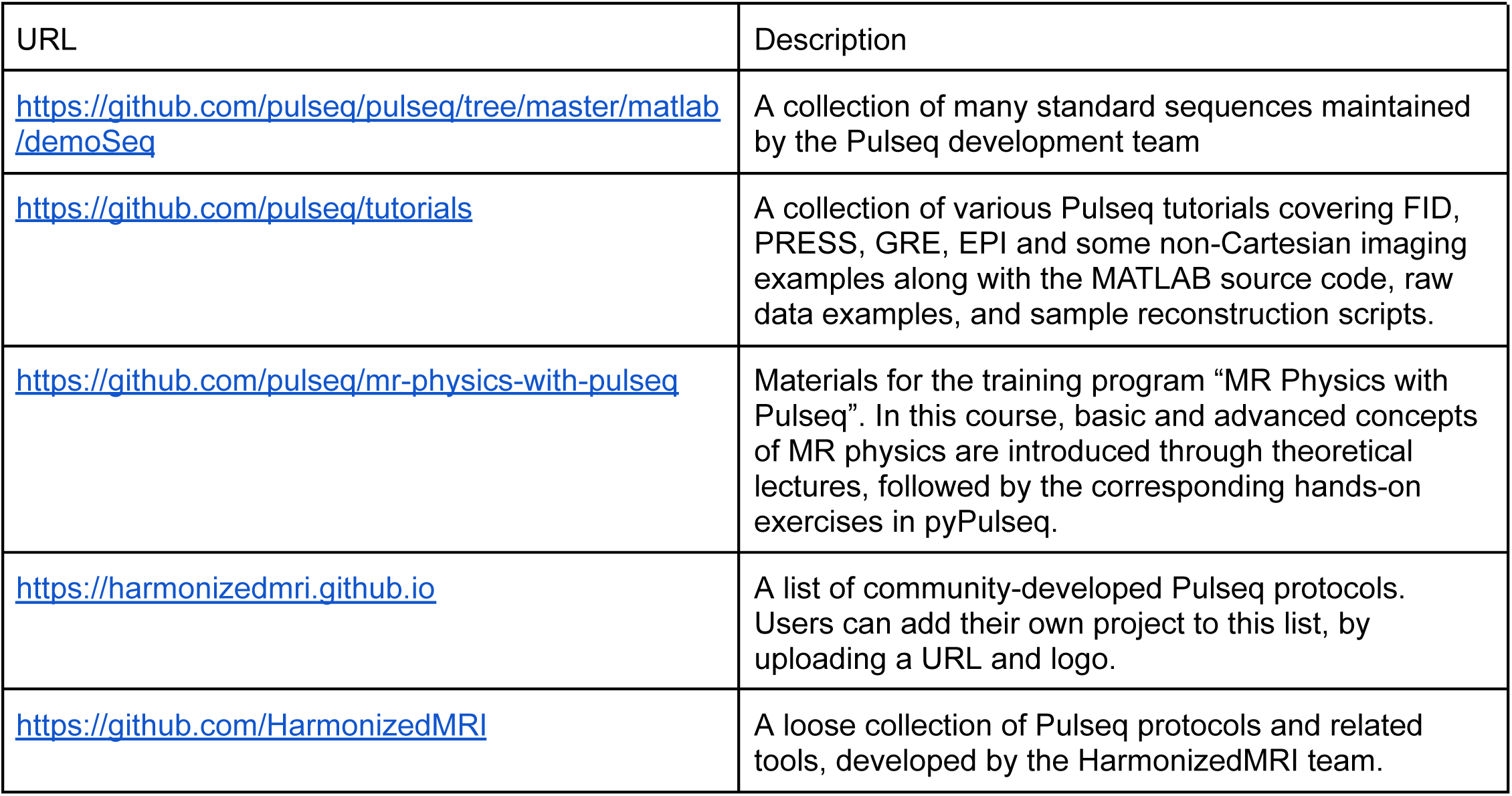
List of web sites containing collections of Pulseq sequence examples.

#### Ensuring compatibility with scanner hardware constraints

It is the responsibility of the sequence designer to ensure that the various RF/gradient/ADC events specified in the .seq file are compatible with the scanner hardware configuration. To aid with this task, Pulseq defines a list of generic hardware parameters that are set by the sequence designer. These include: RF amplitude, dead time, ringdown time, and raster time; gradient amplitude, slew rate, and raster time; ADC dead time and dwell time; and block duration raster time. The Pulseq toolbox includes both internal and user-callable checks to ensure that the sequence object adheres to these settings. For a sequence to run on a specific scanner, its design specifications must be equal to or lower than the actual capabilities of that scanner.

#### Software tools for creating Pulseq files

Apart from the ‘official’ MATLAB toolbox, sequence designers can choose from several alternative ways to create a .seq file. pyPulseq is an open Python implementation of the MATLAB toolbox, and enables sequence creation in environments where it may be difficult to obtain a MATLAB license. Higher-level design tools also exist^7,8^, however, the most common way to create Pulseq (.seq) sequence files is to work directly with the MATLAB^2,5^ and Python^6^ toolboxes.

### Availability of Pulseq interpreters on various MRI vendor platforms

The Pulseq standard is built on a strict separation between the pulse sequence definition (encapsulated in a Pulseq object or .seq file) and the system or software that ‘consumes’ it. This has enabled both the research community and scanner vendors to independently develop various third-party Pulseq tools, including the interpreters that now exist on multiple vendor platforms. Table 2 lists the various interpreters that we are aware of at the time of writing.

**Table 2.**
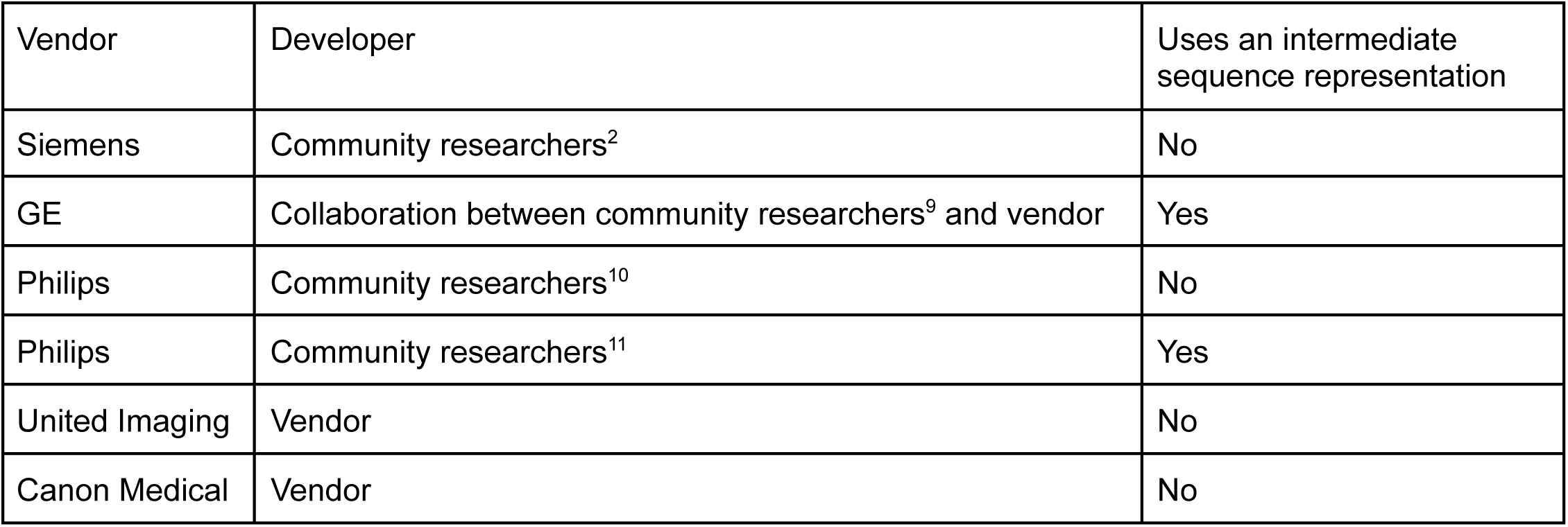
List of available Pulseq interpreters on various human MRI scanner platforms.

These interpreters are at various stages of development, and we encourage the reader to contact the respective developer or their vendor representative for the most up-to-date information. However, some general observations can be made. First, several interpreters have been developed by community researchers using only the vendors’ standard pulse sequence application programming interfaces (APIs)—IDEA for Siemens, EPIC for GE, and PARADISE for Philips. These interpreters are openly available within each vendor’s respective research user community. The fact that they rely solely on widely accessible development tools, combined with their open availability, supports the ongoing development and long-term sustainability of these interpreters. Another important benefit of implementing the interpreters using the vendor APIs is that Pulseq sequences can be plotted and simulated using the standard vendor tools. Figure 5(a) shows an example of this, where our SMS-EPI sequence is simulated and plotted using GE’s native sequence tools (WTools and Pulse View, respectively).

**Figure 5.**
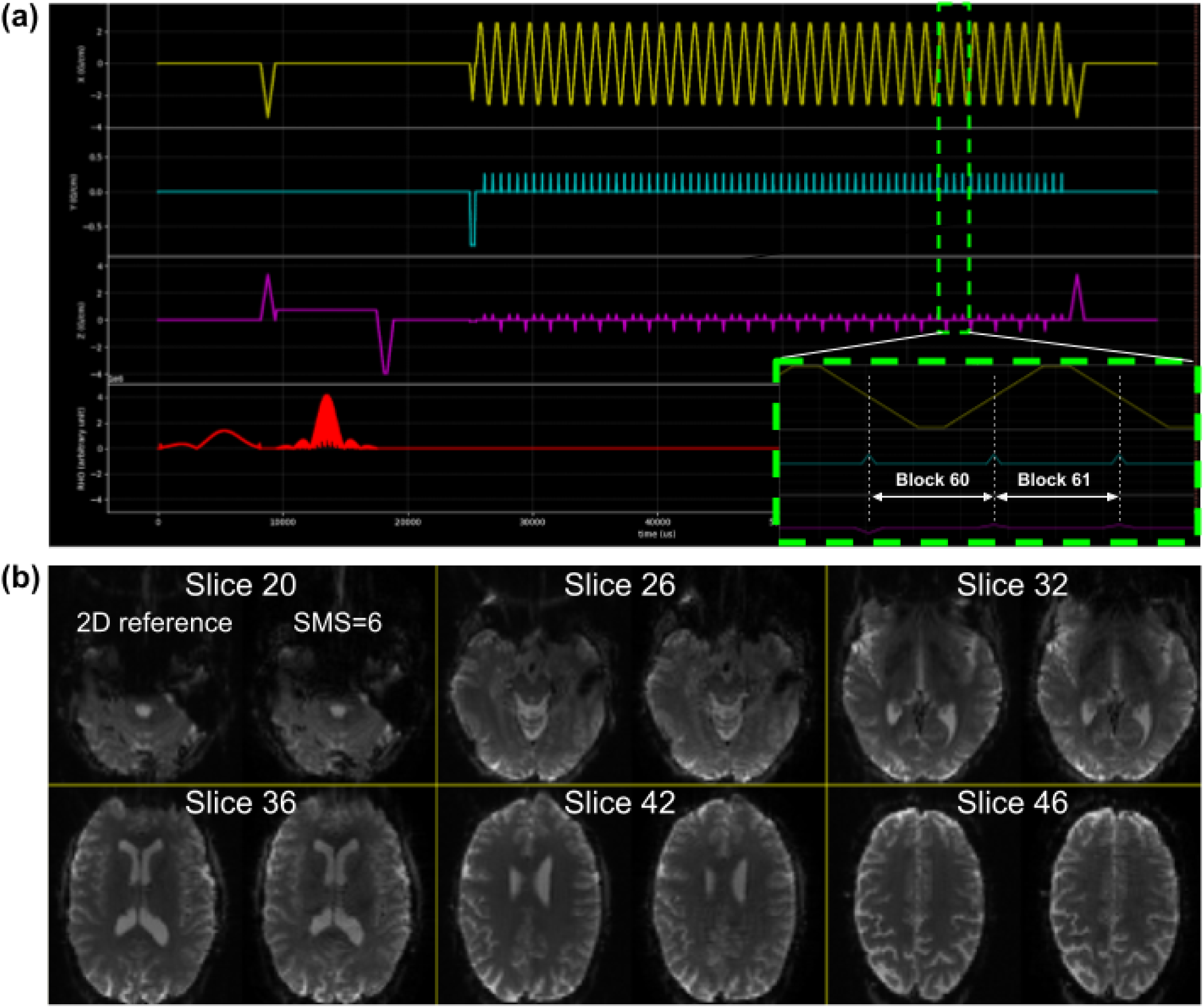
(a) Proposed Pulseq SMS-EPI sequence simulated in GE’s sequence simulator Pulse Studio. The sequence closely matches the ABCD SMS-EPI protocol (CAIPI/SMS factor 6; 2.4 mm iso; FOV = [21.6 21.6 14.4] cm). The inset shows gradient waveform details for two consecutive blocks within the EPI train, (b) Volunteer brain images obtained with the sequence in (a). Six slices are shown: each panel shows a non-SMS (2D) reference EPI image on the left, and the SMS=6 image on the right. The latter was reconstructed with SENSE using BART.

Second, some interpreters require an intermediate sequence representation before execution can proceed. This is the case for GE scanners, where the internal sequence format depends on recognizing repeating subunits, such as a sequence TR or a magnetization preparation segment. Currently, users can convert sequences into this intermediate form manually using openly available tools^12^, or take advantage of the recently introduced Pulserver framework^13^ that automates the conversion process. In addition to simplifying sequence deployment, Pulserver also enables more interactive sequence prescription directly on the scanner console.

Third, vendors have generally demonstrated strong support for the Pulseq platform, as evidenced by the development of vendor-authored interpreters and the substantial technical assistance provided to community developers^13,14^. Vendors appear to recognize the value of supporting their research users through Pulseq, both as a rapid prototyping tool and as a means to facilitate protocol harmonization across sites.

Lastly, as broad vendor platform support continues to emerge—each with its own software and hardware constraints—there will be a growing need for free and open-source software tools that enable researchers to verify the compatibility of a given Pulseq sequence description (e.g., a .seq file) with the available scanner hardware. Developing such tools offers an opportunity for community researchers and developers to actively contribute to the Pulseq project.

### Patient safety and system hardware protection

Deploying an MRI pulse sequence requires careful consideration of both subject safety and hardware protection. Subject safety focuses primarily on limiting RF power deposition (specific absorption rate — SAR) and peripheral nerve stimulation (PNS), while hardware protection involves ensuring the integrity of the RF and gradient subsystems. In sequences developed using vendor-provided pulse programming toolkits (APIs), these safety checks are typically handled either automatically by the scanner or manually by the sequence designer through API-accessible safety routines. The Pulseq interpreters also benefit from these built-in system and API-level safety mechanisms. Additionally, Pulseq allows SAR and PNS to be estimated during the sequence design phase—before the sequence is deployed to the scanner. Here we summarize the various safety checks employed by Pulseq, focusing on the two vendor platforms used so far in this project (Siemens and GE).

#### RF power deposition (SAR)

To our knowledge, all human MRI scanners are equipped with hardware for monitoring RF power deposition. This hardware monitor is always fully engaged, for Pulseq and non-Pulseq sequences alike. In addition, the Siemens and GE interpreters use the API safety routines in the same way as native (product/research) sequences. The Pulseq sequence designer can perform additional checks during the design phase to reduce the chance that the sequence will trip the scanner’s built-in and API SAR checks, such as comparing the nominal RF power to a known reference scan; planning ahead in this way is particularly useful for multi-site studies.

#### Peripheral nerve stimulation (PNS)

The sequence designer can (and should!) model PNS during the design stage using either the SAFE model for Siemens scanners^15–17^, or the nerve impulse response function model for GE scanners^18^. Open MATLAB implementations of these PNS models are freely available on Github. Developing a vendor-neutral, standardized PNS model that can be employed across a broader range of vendors and scanner models is an open problem.

#### Hardware protection

Like pulse sequences implemented with the native API, the Pulseq interpreters rely on API-accessible safety routines to ensure hardware safety. For example, the GE API (EPIC) defines the subroutine minseq to calculate the minimum allowable TR for a given gradient waveform. RF subsystem checks are performed in a similar way. In this way, the vendor does not need to expose any proprietary details about the underlying safety checking algorithms or hardware performance; only the subroutine output (e.g., minimum allowable TR) is needed by the interpreter. This also means that responsibility for hardware protection rests solely on the interpreter, and not on the sequence designer.

### Proposed vendor-neutral, open-source fMRI protocol

#### Pulse sequence implementation

We implemented a BOLD SMS-EPI sequence with parameters that closely match the ABCD protocol^19^: 2.4 mm isotropic resolution, 90x90 matrix size, 60 slices without gaps, SMS factor 6, TR 0.8 s, partial Fourier factor 0.8. We created the various sequence events, and assembled the Pulseq sequence object, using the MATLAB Pulseq toolbox. The MATLAB scripts are freely and openly available on Github^20^, enabling users to modify the acquisition parameters to suit their own needs.

Figure 5(a) shows the timing diagram for one sequence TR, which consists of fat saturation using a minimum-phase Shinnar-Le Roux (SLR) pulse^21^, SMS excitation using SLR sub-pulses with phase offsets following the scheme by Wong^22^, and ramp-sampled EPI readout. To reduce the risk of PNS during the EPI train, we derated the slew rate to ensure that the predicted PNS, based on the nerve impulse response model^18^ stays below 80% of the stimulation limit (normal mode). The sequence is RF-spoiled to suppress steady-state signal contributions from spin- and stimulated-echo pathways^23^.

#### Image reconstruction

To reconstruct images from the acquired raw (k-space) SMS-EPI data, the user is free to choose from several available image reconstruction pipelines. The HarmonizedMRI development team has implemented both 3D SENSE^24^ and slice GRAPPA^25^ SMS-EPI reconstructions in MATLAB that are part of the QA protocol described above. Other community-developed implementations are also available^26–30^. Figure 5(b) shows the results of reconstructing Pulseq SMS-EPI data using 3D SENSE^24^ implemented in BART^29^. Figure 6 demonstrates that our Pulseq protocol achieves similar image contrast and overall image quality as the matched product protocols.

**Figure 6.**
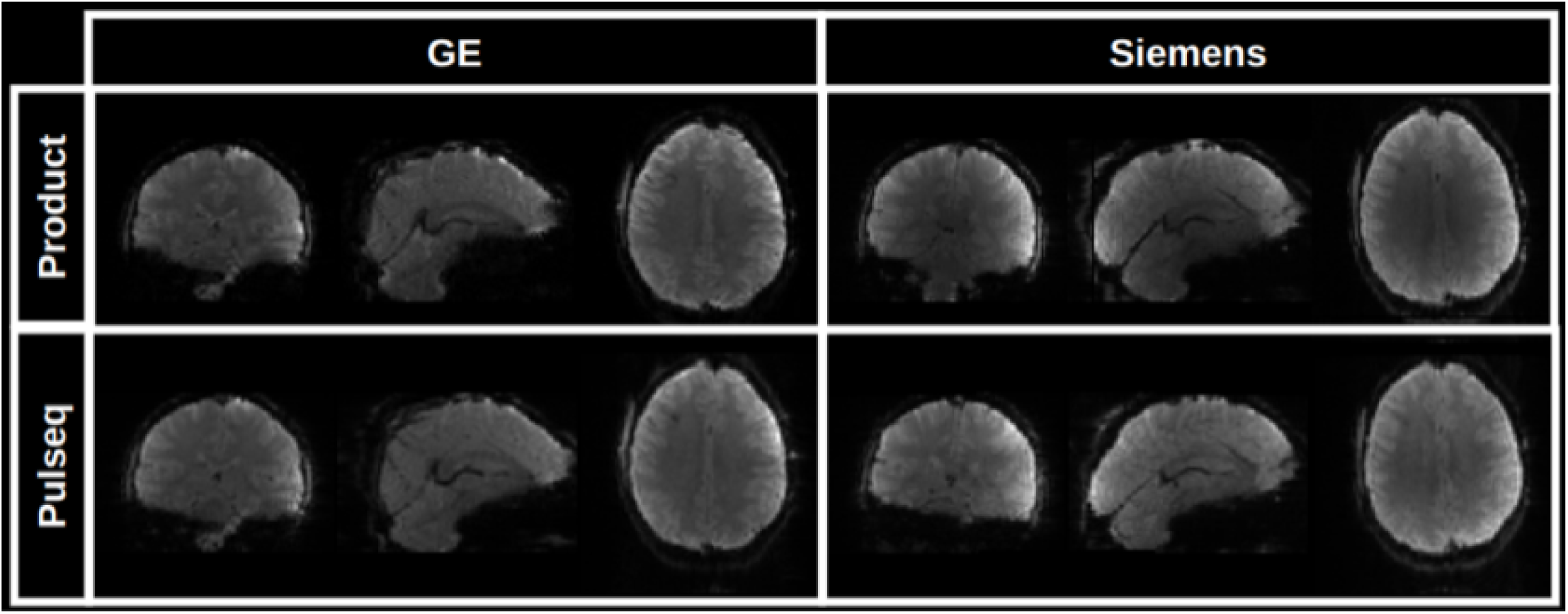
Image comparison between the product (vendor) SMS-EPI protocols (top) and our proposed Pulseq SMS-EPI protocol (bottom). The same subject was scanned on two different scanners (GE and Siemens). The vendor protocols relied on each vendor’s built-in-and closed-image reconstruction algorithm. We obtained the Pulseq images by reconstructing the data offline in MATLAB, using slice GRAPPA.

In our current workflow, image reconstruction is done with slice GRAPPA due to its fast computation speed and robust image quality, and zero-filling partial Fourier reconstruction. Other choices are possible, such as multi-coil reconstructions with spatial regularization, and homodyne PF reconstruction. The extent to which these image reconstruction and post-processing choices impact fMRI reproducibility is an open research question.

### Results: Image quality metrics

To quantitatively assess overall image quality, we analyzed time-series data in phantoms and a volunteer obtained with both the proposed Pulseq SMS-EPI protocol and the vendors’ native (product) protocols on two vendor platforms (GE and Siemens). Figure 7(a) displays the ABCD quality metrics calculated for phantom data acquired on both a GE MR750 and a GE UHP scanner. These values are compared against reference values acquired in the ABCD study. We observe that Pulseq performs favorably against the vendor ABCD protocol. The higher SNR in the ABCD UHP acquisition is partly ascribed to the full ky coverage, as opposed to the fractional ky coverage on the MR750 and in the Pulseq acquisition. As the Pulseq acquisition is the same across scanners, SNR values are more similar across the two scanners as compared to the ABCD protocol.

**Figure 7.**
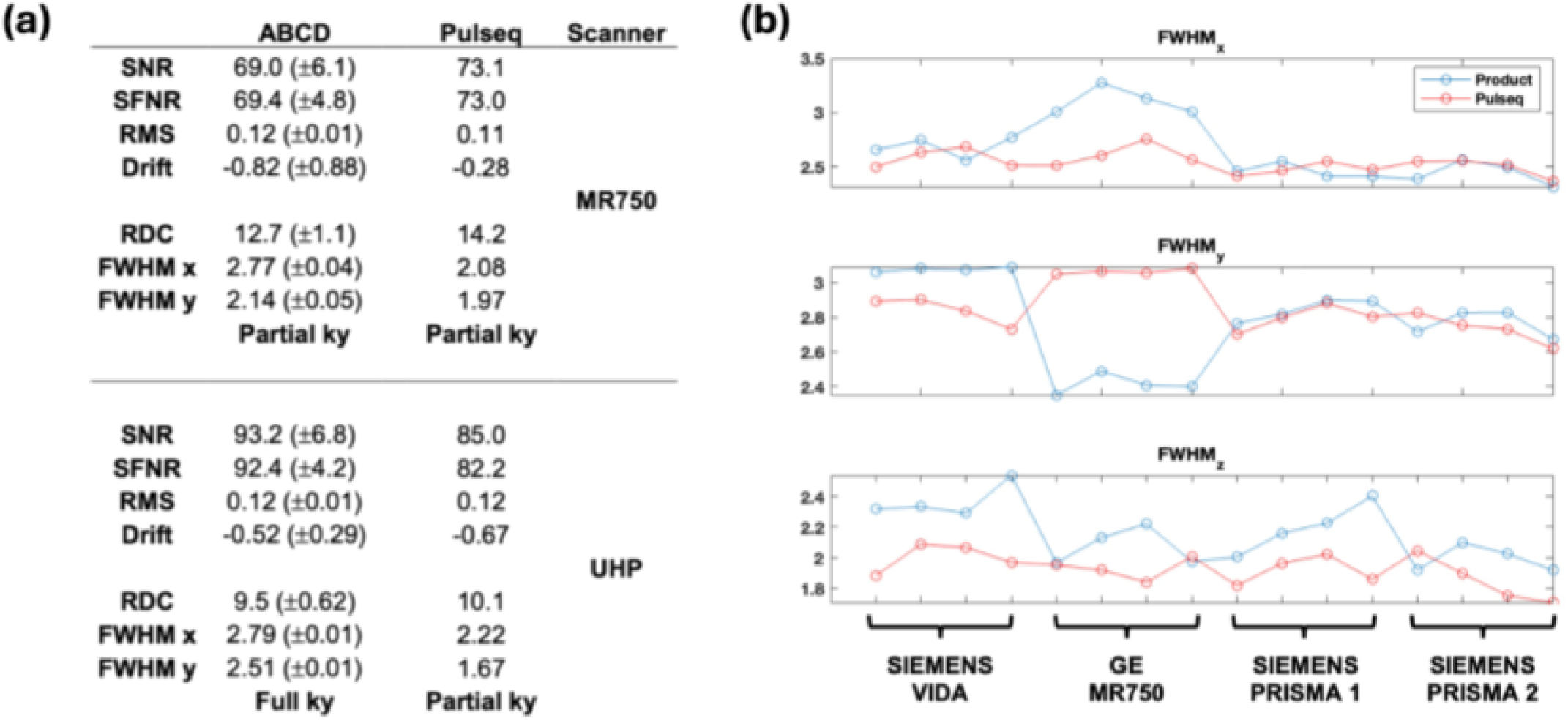
(a) FBIRN phantom metrics for GE MR750 (top) and GE UHP (bottom) MRI scanners, using either ABCD (left) or Pulseq implementation of SMS-EPI. ABCD phantom metrics were taken as the average of multiple phantom scans (MR750: N=120, UHP: N=189). ABCD implementation of SMS used partial ky acquisition on MR750, full on UHP. (b) FWHM metrics for a traveling subject across four scanners, with four resting-state scans on each. SNR: image signal-to-ratio; SFNR: signal fluctuation to noise ratio; RMS: room mean square residual after polynomial fitting; RDS: radius of decorrelation; FWHM: spatial auto-correlation full width half maximum. ABCD: adolescent brain and cognitive development study.

Figure 7(b) plots the FWHM values for the product multiband sequences (blue) vs Pulseq implementation (red) in both GE and Siemens scanners, for resting-state data acquired in a travelling subject, with four repeats at each site. FWHM is a measure of spatial autocorrelation, with lower values being desirable (indicating higher resolution). It is seen that there is reduced variance across scanners and vendors with the Pulseq ABCD acquisition compared to the product ABCD acquisition (ABCD: Adolescent Brain and Cognitive Development study^19^).

### Results: Cross-vendor reproducibility of resting-state functional connectivity

To assess the potential of our Pulseq fMRI protocol for improving cross-vendor reproducibility of resting-state fMRI metrics, we conducted two small traveling subject studies. In the first study, one traveling subject visited 5 separate scanners, and underwent four runs of a five-minute resting-state scan at each site. Figure 8 summarizes the results of this study. It is seen that the Pulseq average connectivity matrix is comparable to the Product connectivity matrix, and that the standard deviation has fewer regions of relatively high cross-scanner variation (though this may be due to subject effects). In other words, this data suggests that harmonizing the acquisition (with Pulseq) as well as the SMS-EPI reconstruction results in more consistent rsFC metrics across these 5 scanners.

**Figure 8.**
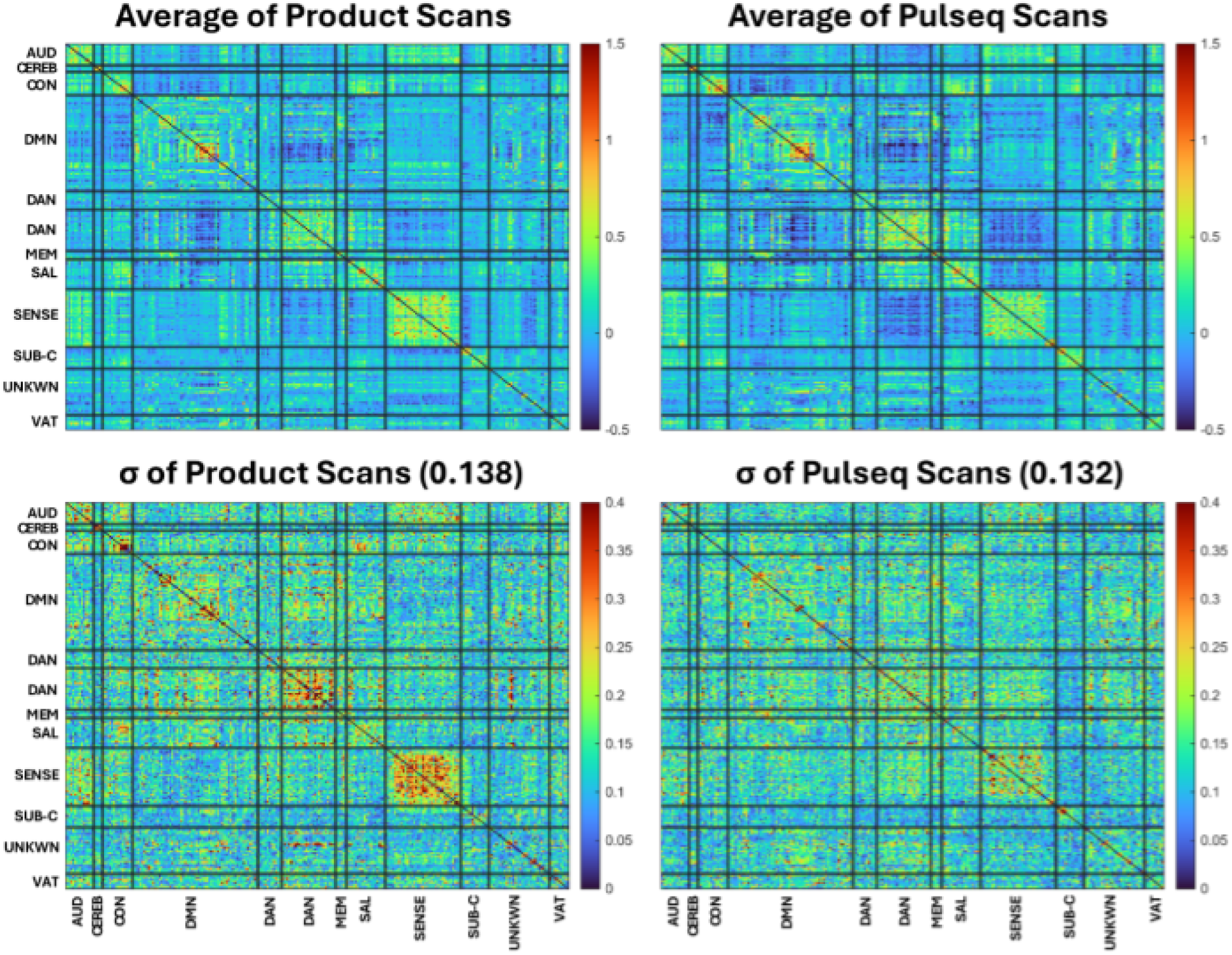
Across-scanner mean (top) and standard deviation (bottom) of resting-state functional connectivity values for Product (left) and Pulseq (right) scans of one subject scanned on 5 different 3T scanners (2 GE; 3 Siemens). The Pulseq protocol produces more consistent rsFC maps (lower standard deviation) for several of the larger networks (e g., sensory and frontoparietal networks).

In the second study, 7 volunteers each underwent two separate scan sessions on different days and on different MRI vendor platforms (Siemens and GE). Each session consisted of a T1 anatomical scan and two 5-minute rest scans, one using the Pulseq protocol and the other the product protocol. In the same session we also acquired 4 repetitions of a block finger-tapping and visual activation task using both Pulseq and product protocols; analysis of the task data is ongoing and will be presented elsewhere. Figure 9 summarizes the resting-state analysis from this study. We observed that the within-subject, cross-scanner resting-state FC matrices tend to be more similar when using the proposed Pulseq protocol compared to the product protocols.

**Figure 9.**
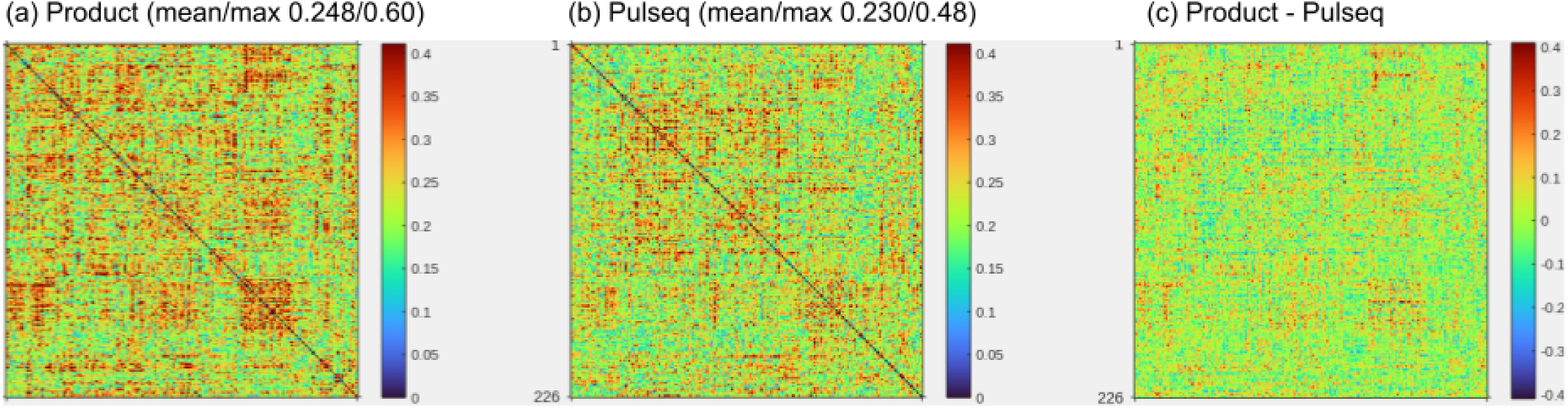
Summary of resting-state SMS-EPI data in 7 volunteers, each scanned on two different 3T scanners (GE MR75O and Siemens Vida), (a-b) Within-subject absolute difference in rsFC maps between the two scanners, averaged over all subjects. The mean (across subjects) absolute difference between the (a) product and (b) Pulseq protocols are shown. The mean and maximum value are indicated. Smaller values (blue and green) are desirable, as it indicates closer agreement between Siemens and GE. The maximum cross-vendor difference is reduced by 20% using Pulseq compared to the product protocols. On average, Pulseq achieves a 7% reduction in cross-vendor rsFC differences. These rsFC difference maps were calculated from images that were smoothed with a 6 mm smoothing kernel as is standard practice, which is likely to mask any differences between the scans, (c) Difference map obtained by subtracting (b) from (a).

#### Functional connectivity methods

Resting-state functional connectivity (rsFC) matrices were calculated for each scan using the CONN toolbox^31^ release 22.v2407^32^ and SPM^33^ (reference) release 12.7771. Functional and anatomical data were preprocessed using CONN’s default preprocessing pipeline^34^ (minus slice time correction) including realignment, outlier detection, direct segmentation and MNI-space normalization, and smoothing with a 6mm kernel. In addition, functional data were denoised using CONN’s standard denoising pipeline^34^ including the regression of potential confounding effects characterized by white matter and CSF timeseries, motion parameters and their first order derivatives, outlier scans, session and scan effects and their first order derivatives, and linear trends within each resting state run, followed by bandpass frequency filtering of the timeseries between 0.008 Hz and 0.09 Hz. The rsFC matrices were calculated using the nodes from the Power functional network atlas^35^.

#### Example usage

To help users get started, we created a complete example showcasing the entire workflow — from data acquisition to generating an image time series. You can explore it here: https://github.com/HarmonizedMRI/SMS-EPI/tree/main/example. The demo includes step-by-step instructions, links to all source code, and access to a sample dataset (available upon request via a Google form).

### Incorporating Pulseq fMRI in your research: Recommendations and additional resources

Thus far, we have focused on the technical implementation of the Pulseq fMRI protocol and presented traveling subject data that demonstrate its scientific value. Based on the information provided in the preceding sections and the associated GitHub repositories, technically proficient users should be able to install the Pulseq SMS-EPI protocol, customize it as needed, and assess its suitability for their research. In this section, we provide additional guidance and recommendations for users at varying levels of engagement and pulse sequence expertise, from initial installation and testing to the planning and execution of multi-site research studies.

#### Quality Assurance (QA) protocols

To ensure correct installation of the proposed Pulseq fMRI protocol and consistent performance across sites, we recommend that the user executes two QA protocols. The first is described in detail in Chen et al.^36^ and involves scanning a uniform ball phantom (preferably FBIRN/EZfMRI) using a set of separate Pulseq sequences available at https://github.com/HarmonizedMRI/qualityAssurance/tree/main. The data is then processed using a set of MATLAB scripts that are also available at that Github site, and that produce performance metrics including signal-to-noise ratio (SNR), temporal SNR, and EPI ghost to signal ratio.

We also recommend calculating quality metrics using the ABCD QC protocol (which is based on the fBIRN protocol) on an fBIRN/EZfMRI phantom, as implemented in the ABCD study. [https://github.com/ABCD-STUDY/FIONA-QC-PHANTOM]. Besides the metrics mentioned above, this protocol also calculates RDC (radius of decorrelation), and FWHM (full-width half max) for each spatial axis; both of which can help detect issues in image quality.

For both QA protocols, we further recommend that the user compare their results against the product protocol on their scanner, and against normative values that we provide.

#### Getting technical help and connecting with other users

Our open fMRI protocol is part of a broader ecosystem of open-source software for sequence design and image reconstruction, and vendor-specific Pulseq interpreters that are typically only available within each vendor’s research user community. As such it can be difficult for the resource user to know where to look for guidance on specific components of the proposed fMRI workflow, particularly in a multi-vendor, multi-site setting. Here we provide a few resources to help orient the user. For help getting started with the proposed SMS-EPI fMRI protocol that is the focus of this article, see https://github.com/HarmonizedMRI/Functional.

#### General help with Pulseq

The formal Pulseq specification and the source code for the MATLAB toolbox are available at https://github.com/pulseq/pulseq. That site also contains an active discussion forum and a bug report forum; these are named ‘Discussions’ and ‘Issues’, respectively, following the usual Github platform convention. In addition, several recorded Pulseq talks are available online.

#### Pulseq user group meetings

Beginning in September 2025, we will host a regular virtual meeting for the Pulseq user community, to provide technical help in an ‘office hour’ format, and give community researchers an opportunity to present and discuss their work in an informal setting. This meeting is open to everyone -- a registration link is provided on the project home page (https://github.com/HarmonizedMRI/Functional).

#### Direct help from members of the HarmonizedMRI study team

We are making a fee-based service package available to funded fMRI studies that includes assistance with protocol installation and QA, protocol maintenance across scanner software upgrades, and consultation. We suggest that PIs budget for this service package in their grant proposals; suggested template text to include in the NIH Budget Justification section is described below.

#### Writing a multi-site research study grant proposal

To assist Principal Investigators (PIs) in integrating the proposed fMRI protocol into future multi-site research studies, we offer example/template text suitable for inclusion in various sections of a grant proposal^37^. While the template is primarily designed for NIH applications, it can be adapted to meet the requirements of other funding agencies. Users are encouraged to tailor the text as needed to align with their specific project goals and proposal style. The templates include the following:

- Scientific and technical rationale for using Pulseq (Research Plan section)
- QA report from all study sites (Research Plan or Facilities & Resources)
- Budget justification section:

- Funding for FBIRN phantom (for QA protocol)
- Tiered support subscription service (optional)
- Human subjects protection section (Pulseq-specific statements regarding safety)
- Letter of support from MRI scanner manufacturer(s) (optional)

While optional, we recommend including letters of support from the vendors. From the vendors’ standpoint, executing a Pulseq sequence is equivalent to running any custom pulse sequence developed by a research user through the vendor’s programming APIs. Consequently, each scanner participating in a proposed multi-site study must have an active research license. Provided this requirement is met, vendors should be able to furnish a letter of support granting permission to run the Pulseq fMRI protocol, confirming access to the standard technical support channels available to research users on that platform, and—ideally—endorsing the scientific rationale of both the Pulseq fMRI protocol and the broader Pulseq initiative. To facilitate this process, we provide a sample letter of support that incorporates these elements, that users are free to adapt to their own needs.

### Preparing the IRB protocol

#### Pulseq-specific IRB protocol

As noted above, Pulseq sequences are implemented using the standard pulse sequence programming environments provided by each vendor (e.g., EPIC for GE, IDEA for Siemens) and are subject to the same safety checks and hardware constraints as any other research sequence. We nevertheless recommend preparing a dedicated IRB protocol specifically for general scanning with Pulseq sequences. This protocol only needs to be created once and can significantly reduce administrative burden by eliminating the need to repeatedly explain the nature and risks of Pulseq in each new study. The Pulseq IRB protocol should be used in conjunction with the study-specific IRB protocol, and participants should provide informed consent for both.

#### Joining the University of Michigan multi-site (single) IRB protocol

An alternative for researchers based in the United States is to cede IRB oversight to the University of Michigan. This eliminates the need to prepare a site-specific Pulseq IRB application, and further encourages scientific collaboration and data sharing between research sites. The U of Michigan multi-site (“single”) IRB protocol covers general Pulseq scanning including both functional and structural MRI, and is currently in preparation. We share the scientific protocol text publicly^38^, which can also be used as a template for preparing a site-specific Pulseq IRB application. The U of Michigan Pulseq IRB protocol should be used in conjunction with the study-specific (typically local) IRB protocol, and participants should provide informed consent for both.

## Discussion

The proposed Pulseq fMRI protocol enables identical sequence execution and image reconstruction across multiple scanner platforms, providing highly reproducible conditions for multi-site fMRI research. While this approach minimizes variability, some inter-site differences inevitably remain—for example, variations in receive coil arrays and RF transmit coil configurations. Another source of variability is the B0 shimming protocols employed by different vendors; however, recent efforts by our group^39^ and others^40^ have begun to address this issue. A more subtle factor is the accuracy of gradient trajectories, which can be influenced by differences in gradient hardware performance and vendor-specific eddy current compensation routines. Developing a standardized method for measuring gradient trajectories—and evaluating their impact on the quality of the derived functional maps/measures—remains an area for future research.

As currently implemented, our Pulseq fMRI protocol only allows limited prescription-time flexibility. While the field-of-view can be freely moved and rotated, most other acquisition parameters are fixed including the matrix size and SMS acceleration factor; modifying these parameters requires editing the MATLAB script and creating a new .seq file. In some imaging settings, such as during initial image quality assessment and protocol optimization, it can be beneficial to allow a more interactive sequence prescription, ideally using the vendor’s built-in user interface (UI). Pulserver^13^ is a recent effort to provide this functionality, but has so far only been demonstrated on one vendor platform. Another approach is the gammaStar framework^41^, which uses a dedicated user interface and sequence generation backend. Ongoing work is exploring the use of these frameworks to support interactive, vendor-neutral MRI protocol development.

At present, our fMRI protocol includes offline image reconstruction, which is generally sufficient for functional imaging studies. However, clinical research settings can benefit from online (on-the-host) image reconstruction and display similar to the built-in sequences, e.g., as an immediate image quality check and to integrate directly with the standard DICOM image workflow and storage. To our knowledge, most MRI vendors now offer containerized, online image reconstruction environments that allow research users to plug their own custom algorithms into the scanner’s image reconstruction and display pipeline. Together with a more interactive sequence prescription as just described, these containerized environments will enable vendor-neutral (f)MRI protocols that have a similar ‘look and feel’ as built-in protocols.

The vendor interpreters we have tested so far (Siemens, GE) do not yet allow real-time sequence updates, as would be necessary for, e.g., navigator-based prospective motion correction. While existing fMRI protocols in routine use rely on retrospective image co-registration, prospective motion correction may have certain advantages, such as reduced slice interpolation artifacts. To our knowledge, the vendors’ real-time sequence update mechanisms are available to their research users (including the Pulseq interpreter developers), making it possible to modify the Pulseq interpreters accordingly. Implementing and testing real-time sequence updates for Pulseq remains an area for future research.

The current SMS-EPI acquisition and reconstruction methods we offer are based on well-established approaches, but there remains potential for further improvement. We anticipate—and actively encourage—the research community to use this implementation as a foundation or benchmark for developing improved fMRI protocols.

## Conclusion

The proposed fMRI protocol harmonizes all software-related aspects of an SMS-EPI protocol, from data acquisition to image reconstruction. The protocol is fully open source, and has so far been tested on two vendor platforms (Siemens and GE) with more expected in the near future. We provide resources to help researchers draft grant and IRB applications that use the proposed Pulseq fMRI protocol, and customer support for sites without a dedicated MRI physicist.

## Back matter

### Data and code availability

**Code for Pulseq sequence file generation and image reconstruction:** The code used in this manuscript to create the Pulseq sequence files, and reconstruct the data to form time-series images, is publicly available. The home page for this project, https://github.com/HarmonizedMRI/Functional, contains links to this code as well as a general overview of the project. A complete, tutorial-style, example of using this code is available at https://github.com/HarmonizedMRI/SMS-EPI/tree/main/example.

**Pulseq interpreters:** Source code for the vendor-specific GE and Siemens Pulseq interpreters used in this manuscript is freely available within the respective vendor research user communities. GE research users can request access to the EPIC source code by navigating to https://github.com/HarmonizedMRI/SequenceExamples-GE/. Siemens research users can acquire the Pulseq Interpreter under the conditions of the Customer-to-Customer Partnership (C^2^P, a.k.a. C2P) program, either electronically via the TeamPlay web service or by contacting the authors at.

**Example text for NIH grant and IRB applications:** The project home page (https://github.com/HarmonizedMRI/Functional) contains a link to the example grant and IRB text templates mentioned in this manuscript. This text is provided at no cost and without constraints. Users therefore are free to adapt the text to suit their own needs, and to use it for any purpose.

**Example volunteer brain fMRI data set:** The data set used in the example at https://github.com/HarmonizedMRI/SMS-EPI/tree/main/example is available upon request (via a Google form, available at that site). The data set consists of raw, multi-coil, SMS-EPI k-space data. Our current IRB does not permit publicly sharing raw k-space data since such data cannot be de-faced or otherwise made fully anonymous. The user must agree to not attempt to identify any study participants.

### Funding sources

Funding for this project is provided by NIH grant number U24-NS120056.

### Conflict of interest statement

The authors have no conflicts of interest to disclose in connection with the content of this manuscript.

## Data Availability

The example data set used in the protocol workflow sample at https://github.com/HarmonizedMRI/SMS-EPI/tree/main/example is available upon request, via an online submission form on that site.

## Notes

### Competing Interest Statement

The authors have declared no competing interest.

### Funding Statement

Funding for this project was provided by NIH grant number U24-NS120056.

### Author Declarations

This study was approved by the University of Michigan Institutional Review Board. All subjects provided informed consent.

